# Transmission of Acute Respiratory Infections during Aerosol Generating Medical Procedures (AGMPs): An updated review

**DOI:** 10.1101/2021.11.05.21265762

**Authors:** Jenine Leal, Mark Hofmeister, Liza Mastikhina, John Taplin, Joyce Li, Brenlea Farkas, Laura Dowsett, Tom Noseworthy, Fiona Clement

**Affiliations:** Department of Community Health Sciences, University of Calgary, Calgary, Alberta Canada; Department of Microbiology, Immunology and Infectious Diseases, University of Calgary, Calgary, Alberta Canada; O’Brien Institute for Public Health, University of Calgary, Calgary, Alberta Canada; Infection Prevention and Control, Alberta Health Services, Alberta Canada

**Author notes:** Corresponding Author: Jenine Leal, PhD, Foothills Medical Centre, 801 - South Tower, 3031 Hospital Drive NW, Calgary, AB T2N 2T8. Mark Hofmeister, 3D10, 3280 Hospital Drive NW, Calgary, AB T2N 4Z6. Liza Mastikhina, 3D10, 3280 Hospital Drive NW, Calgary, AB T2N 4Z6. John Taplin, 3D10, 3280 Hospital Drive NW, Calgary, AB T2N 4Z6. Tom Noseworthy, 3D10, 3280 Hospital Drive NW, Calgary, AB T2N 4Z6. Joyce Li, 3D10, 3280 Hospital Drive NW, Calgary, AB T2N 4Z6. Brenlea Farkas, 3D10, 3280 Hospital Drive NW, Calgary, AB T2N 4Z6. Laura Dowsett, 3D10, 3280 Hospital Drive NW, Calgary, AB T2N 4Z6. Fiona Clement, 3D10, 3280 Hospital Drive NW, Calgary, AB T2N 4Z6. **Author Contributions Statement:** *Conceptualization of study:* JLeal. Methodology: JLeal, FClement, LDowsett, MHofmeister, LMastikhina, JTaplin, JLi, BFarkas. *Supervision:* FClement, TNoseworthy, JLeal. *Analysis:* LDowsett, MHofmeister, LMastikhina, JTaplin, JLi, BFarkas. *Writing of original draft:* JLeal, LDowsett, MHofmeister, LMastikhina, JTaplin, JLi, BFarkas. *Reviewing and editing draft:* FClement, TNoseworthy, MHofmeister, LDowsett, LMastikhina, JTaplin, JLi, BFarkas.

**Keywords:** aerosol-generating procedures, acute respiratory infection, healthcare worker

## Abstract

1

**Objectives:** To review the literature from 2011 until March 31^st^, 2020 to identify the risk of transmission of ARIs to healthcare workers caring for patients undergoing AGMPs compared with the risk of transmission when caring for patients not undergoing AGMPs.

**Results:** Only two prospective cohort studies were identified meeting inclusion criteria. One found that performance or assistance with AGMP during the previous week was significantly associated with symptomatic influenza (adjusted OR: 2.29, 95% CI: 1.3 to 4.2). The second study found that performance of AGMP was significantly associated with clinical respiratory infections (RR 2.9, 95% CI 1.42-5.87, p<0.01), laboratory-confirmed virus or bacteria (RR 2.9, 95% CI 1.37-6.22, p=0.01), and laboratory-confirmed virus (RR 3.3, 95% CI 1.01-11.02, p=0.05). Further evidence is needed regarding what constitutes an AGMP and the risk of ARI transmission during presumed AGMPs. Organizations need to interpret these findings with caution when establishing AGMP lists requiring airborne precautions.

## 2 INTRODUCTION

Severe acute respiratory syndrome coronavirus 2 (SARS-CoV-2) pneumonia (COVID-19) was first identified in December 2019 in Wuhan, China and has since spread globally. Available evidence suggests the primary route of human-to-human transmission of the SARS-CoV-2 virus is through large respiratory droplets and contact routes.[1, 2] This is supported by studies demonstrating transmission being highest within households,[2, 3] the absence of identified transmission on aircraft,[4] limited reports of significant outbreaks in staff of COVID-19 treatment centres not using airborne precautions and N95 respirators.[5-7] Some studies have measured viral RNA in the air and air vents at enough distance from patients suggesting that aerosol transmission may be possible; however, whether the viral RNA in air represents living virus remains unknown.[8-13]

Certain medical procedures used in the treatment of acute respiratory infection (ARI) may increase the risk of aerosol generation from a patient above that of natural processes (coughing etc.).[14] The World Health Organization (WHO) and the Centers for Disease Control and Prevention (CDC) have identified several potentially aerosol generating procedures, which necessitate the use of airborne precautions such as N95 or higher respirators, in addition to gowns, gloves and eye protection along with airborne infection isolation room.[15, 16] However, there has been very limited research on which medical procedures should be considered aerosol-generating and the evidence defining relative risk of presumed AGMPs is insubstantial.[14, 17]

A systematic review on AGMPs and the associated risk of transmission of ARIs by Tran et al. (2012) identified that some procedures potentially capable of generating aerosols were associated with increased risk of SARS transmission or were a risk factor for transmission.[18] Considering the COVID-19 pandemic, updated evidence is required on transmission of ARIs to HCWs due to aerosolized particles.

## 3 MAIN TEXT

### 3.1 Literature Search

The search strategy by Tran et al. was updated by a research librarian to capture studies in MEDLINE, Embase, Cochrane Database of Systematic Reviews, Cochrane Central Register of Controlled Trials, and CINAHL published from 2011 to March 31^st^, 2020.[18] Terms related to AGMPs were combined with relevant disease terms and searched as text words in titles and abstracts and as MeSH subject headings, when applicable. The search excluded case reports, animal studies, conference abstracts, editorials, and letters. The full search strategy is reported in Additional File 1.

### 3.2 Literature Selection

PRIMSA guidelines were followed (Additional File 2).[19] Abstracts identified were screened in duplicate and all included abstracts proceeded to full-text review. Full-text publications were screened in duplicate with discrepancies between reviewers resolved through consensus. Eligibility criteria included: 1) study population of HCWs caring for patients with ARIs, including but not limited to COVID-19, SARS, MERS, influenza, 2) the intervention was the provision of care to patients undergoing AGMPs, 3) comparator was the provision of care to patients not undergoing AGMPs, 4) the outcome was the risk of transmission of ARIs from patients to HCWs and 5) the study design was a randomized controlled trial or non-randomized comparative study. Publications were excluded if they were not published in English or French.

### 3.3 Data Extraction & Analysis

For all included studies, year of publication, country, study design, population, aerosolizing procedure, period of evaluation, assessment of training and protective equipment, and number of exposed and non-exposed cases were extracted in duplicate using standardized data extraction forms. Discrepancies between reviewers during data extraction were resolved through consensus. Results of these studies were narratively synthesized.

## 3.4 Results

Searches of electronic databases yielded 7,299 records. The full texts of 59 records were screened, of which two records met inclusion/exclusion criteria (**Error! Reference source not found**.) (Table of excluded studies, in Additional File 3).

**Fig 1.** Flow Chart of Included Studies.

Two prospective cohort studies were identified meeting inclusion criteria (Table 1).

**Table 1.**
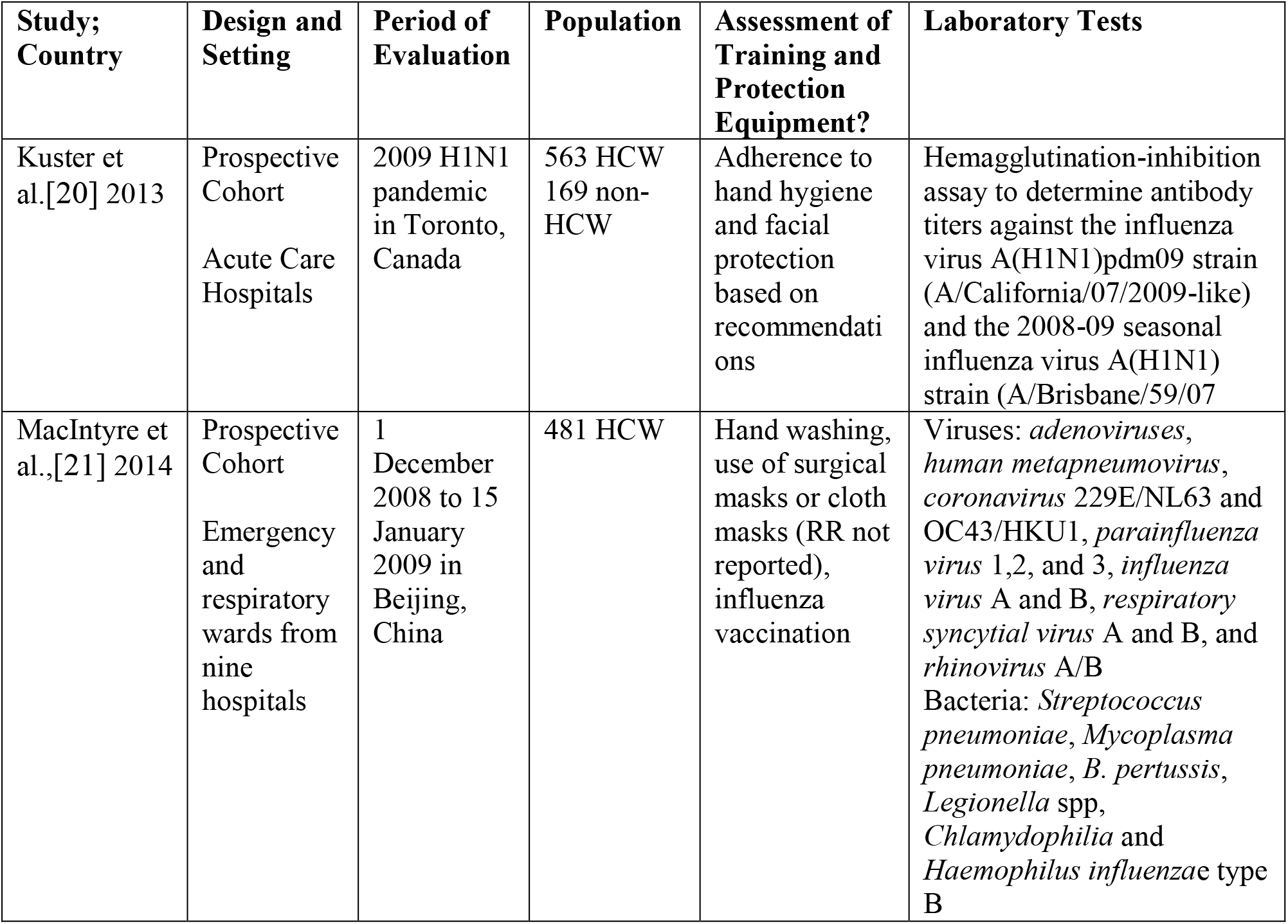
Characteristics of studies included

The primary purpose of the study conducted in Toronto was to examine the epidemiology of infection caused by influenza virus A(H1N1)pdm 09 among HCWs and other working adults in Canada.[20] The authors secondarily determined risk factors for laboratory-confirmed symptomatic influenza among HCWs. An AGMP was defined as any one of the following: administration of nebulized therapy or humidified oxygen at >40%, use of bag-valve mask, manual ventilation, non-invasive ventilation, open airway suctioning, bronchoscopy or other upper airway endoscopy, tracheostomy, endotracheal intubation, cardiopulmonary resuscitation, oscillatory ventilation, or any procedure that involved manipulation of open ventilatory tubing in a mechanically ventilated patient, or sputum induction or other deliberate induction of coughing.[20] In multivariate analysis adjusted for receipt of vaccine and dynamics of pandemic waves, Kuster et al. (2013)[20] found that performance or assistance with AGMP during the previous week was significantly associated with symptomatic influenza (adjusted OR: 2.29, 95% CI: 1.3 to 4.2).[20]

In the second study, MacIntyre et al. (2014) prospectively studied 481 HCWs from emergency and respiratory wards from nine hospitals in Beijing China. Healthcare workers were subjects of a control group in a larger cluster randomized control trial (RCT) comparing fit-tested and non-fit tested N95 respirators to medical masks to prevent respiratory viral infection in HCWs. High risk procedures were defined as: provision of nebulizer medications, suctioning, intubation, AGMPs, and chest physiotherapy.[21] MacIntyre et al. (2014)[21] measured transmission of clinical respiratory infections defined as: presence of two or more respiratory symptoms; or, one respiratory symptom and one or more systemic symptoms. Laboratory-confirmation of viruses and bacteria included are listed in Table 1. Fifty-six (11.6%) HCWs performed at least one high risk procedure during the study, with the most common activity being airway suctioning (66%, 37/56). Using Poisson regression, MacIntyre et al. (2014)[21] found that performance of AGMP was significantly associated with clinical respiratory infections (RR 2.9, 95% CI 1.42-5.87, p<0.01), laboratory-confirmed virus or bacteria (RR 2.9, 95% CI 1.37-6.22, p=0.01), and laboratory-confirmed virus (RR 3.3, 95% CI 1.01-11.02, p=0.05).

## 3.5 Discussion

Two studies were identified in this update of the systematic review conducted by Tran et al.[18] Both additional studies concluded that the performance of AGMPs significantly increased risk of ARI transmission to HCWs.

In both studies, there was a lack of power related to small number of cases of ARIs in the HCW groups and AGMPs were combined. Hence, risk could not be attributed to individual procedures as it was in the study by Tran et al. The study by Kuster et al (2013) was further limited by the potential of: enrollment bias with increased participation by individuals who perceive having a higher risk for influenza infection; recall bias with ill participants potentially reporting risk factors more accurately than people who did not develop the illness; and a Hawthorne effect whereby participants may have altered their behaviour (e.g. increased adherence to protective measures) as a result of participating in the study.[20] Although the participants in the MacIntyre et al. study were part of a control group of a cluster RCT, similar limitations such as recall bias may persist.[21]

Interpretation of these findings is complicated by variation across jurisdictions in clinical care; quantity and type of AGMPs performed, infection prevention and control (IPC) practices, availability of personal protective equipment (PPE); type of PPE; and individual use and fit of PPE for each procedure. In addition to these sources of variation, validity of estimates from meta-analysis in the Tran et al. study was threatened by the very low quality of evidence available. Given this, the interpretation and practical application of the study findings is difficult.

Despite the limited and insubstantial evidence on the types of procedures that constitute an AGMP and the risk associated with presumed AGMPs, many organizations and medical societies have taken a precautionary approach and adopted the full or partial list of procedures identified by Tran et al. (2012).[1, 16, 22, 23] The Infectious Disease Society of America identified that there was no direct evidence of AGMPs and rates of COVID-19 infection among healthcare workers and that despite the very low quality and indirect evidence, they placed a high value on avoiding serious harms to exposed health care personnel.[23] They present the CDC and WHO’s variable lists and rank the procedures from Tran et al. from highest odds ratio to lowest, despite many of the 95% confidence intervals crossing one and not being statistically significant.[23] A more transparent approach would be to present the results highlighting the procedures that have consistently demonstrated higher risk (e.g. Tracheal intubation). With the uncertainty and caution surrounding risk of transmission of SARS-CoV-2 among HCWs, other procedures have been proposed as potential AGMPs despite limited to no studies pertaining to their risk.[24-26] In our jurisdiction, a similar list of procedures was proposed by various clinical groups and societies. This list was reviewed by an expert working group made up of IPC physicians and professionals, workplace health and safety physicians, epidemiologists and respiratory therapists. Where decisions to exclude procedures as AGMPs differed from national/international clinical societies, engagement with these local groups occurred to establish consensus.[27] Other jurisdictions have employed similar processes to develop lists of AGMPs.[28] The result of including or excluding procedures as AGMPs using this approach will differ across jurisdictions depending on IPC guidelines and procedures, perceived risk of transmission and infection, and availability of PPE.

In the context of COVID-19, the evidence for HCW exposure through AGMPs is also limited and the generalizability of these findings to the current COVID-19 outbreak is unclear.[29] In a case control study among HCWs with acute respiratory symptoms working in the designated hospital of Wuhan University, HCW risk of developing COVID-19 was higher in areas where AGMPs were performed (ICUs, respiratory wards, infection department and surgical department).[30] However, this risk was nullified when exposure was stratified by specific AGMPs (e.g. tracheal tube removal, sputum suction, fiber bronchoscopy). Additionally, HCW risk increased substantially on these units by self reported inadequate hand hygiene practices.[30] An epidemiological investigation was conducted of a patient diagnosed with COVID-19 after being nursed in an open ward with 10 other patients for 35 hours and being on high flow oxygen for 18.5 hours before being transferred to an airborne isolation room. Thirty-two of 71 staff developed respiratory symptoms but were all negative for SARS-CoV-2.[6] In another study, 41 HCWs were exposed to AGMPs for at least 10 minutes at a distance less than 2 metres during the care of a retrospectively diagnosed COVID-19 patient in an ICU setting.[7] The AGMPs included endotracheal intubation, extubation, noninvasive ventilation and exposure to aerosols in an open circuit. A majority (85%) of exposed HCWs wore surgical masks, rather than N95 respirators, and followed standard contact and droplet precautions. None of the exposed HCWs developed symptoms and all tests for SARS-CoV-2 were negative. [7]

## 3.6 Conclusion

Like the Tran et al. review, this update finds the presence of a significant research gap which is further compounded by the lack of precision in the literature regarding the definition for AGMPs.[22] Further evidence is needed regarding the probability and types of infections following exposure, by type of procedure and PPE. Multidisciplinary teams should strive to use robust epidemiologic, engineering and aerodynamics or aerobiology study designs to identify associations between viral particles expelled and collected from aerosols generated by these procedures with viable virus capable of infecting susceptible hosts. This would enhance the transferability of lessons learnt from AGMPs, whether from SARS, H1N1 or COVID-19 and prepare us for the next wave of COVID-19 or future pandemics of a respiratory nature.

## 4 LIMITATIONS

This review has limitations given the expected rapid turnaround for completion. We were not able to search the grey literature for additional publications and we did not apply the GRADE assessment to the two studies. Individual AGMPs could not be delineated from the grouped analysis in the studies and therefore risk could not be assigned to individual procedures. The search strategy included terms and key words originally identified in the Tran et al. study; however, since the search, clinical societies have proposed the inclusion of other procedures, which were not included in the original search. However, many of these recommendations have not been substantiated by existing evidence so we do not think we have missed a significant portion of the literature.

## Data Availability

All datasets supporting the conclusions of this article are included within the article

## LIST OF ABBREVIATIONS

AGMPs: aerosol-generating medical procedures
ARIs: acute respiratory infections
CDC: Center for Disease Control and Prevention
HCWs: healthcare workers
ICUs: intensive care units
IPC: infection prevention and control
OR: odds ratio
PPE: personal protective equipment
RCT: randomized control trial
RR: relative risk
SARS: severe acute respiratory syndrome WHO World Health Organization

## 5 DECLARATIONS

### Ethics approval and consent to participate

Not applicable

### Availability of data and material

Not applicable. All data and material available in the manuscript.

## Acknowledgements

We would like to Dr. John Conly, the senior author of the original systematic review for reviewing and commenting on this update.

## Consent for publication

Not applicable

## SUPPORTING INFORMATION CAPTIONS

A1 Appendix.docx: Additional File 1. Complete Search Strategy

A2 Appendix.doc: Additional File 2. PRISMA 2009 Checklist

A3 Appendix.docx: Additional File 3. Table of Excluded Articles

