## Supplementary material for "Transmission of Acute Respiratory Infections during Aerosol Generating Medical Procedures (AGMPs): An updated review": A1 Appendix

**Additional File 1. Complete Search Strategy**

**MEDLINE Final March 31 2020**

1. exp Positive-Pressure Respiration/
2. exp High-Frequency Ventilation/
3. exp Ventilators, Mechanical/
4. Ventilation/
5. exp Intubation, Intratracheal/
6. Suction/
7. Tracheostomy/
8. Bronchoscopy/
9. Thoracostomy/
10. exp "Nebulizers and Vaporizers"/
11. Sputum/
12. Oxygen Inhalation Therapy/
13. Autopsy/
14. exp Respiratory Function Tests/
15. exp Spirometry/
16. exp Cardiopulmonary Resuscitation/
17. exp Respiration, Artificial/
18. exp Breathing Exercises/
19. Physical Therapy Modalities/ and Thorax/
20. (ventilation or ventilator or ventilating or ventilatory).ti,ab.
21. (respirator or respirators or respirat* support or respirat* care).ti,ab.
22. (intubation or intubated or extubation or extubated).ti,ab.
23. ((respiratory or airway or air way or open) adj3 suction*).ti,ab.
24. (nebulize* or nebulise* or aerosolize* or aerosolise*).ti,ab.
25. heat moisture exchange*.ti,ab.
26. (bronchoscopy or tracheostomy or thoracostomy).ti,ab.
27. (chest adj3 physiotherapy).ti,ab.
28. (sputum adj3 (induction or inducing)).ti,ab.
29. oxygen therap*.ti,ab.

30. (lung function test* or pulmonary function test*).ti,ab.
31. ((continuous or bilevel) adj2 (positive airway or positive pressure)).ti,ab.
32. (cardiopulmonary resuscitation or artificial resuscitation or artificial respiration).ti,ab.
33. (autopsy adj3 lung tissue*).ti,ab.
34. (bag adj1 mask*).ti,ab.
35. (ambu bag* or bag valve* or bvm).ti,ab.
36. (manual resuscitator* or self-inflating bag*).ti,ab.
37. ((nasopharyn* or nasal or nose or np or oropharyn* or OP) adj3 (culture* or smear* or specimen* or swab*)).ti,ab.
38. 1 or 2 or 3 or 4 or 5 or 6 or 7 or 8 or 9 or 10 or 11 or 12 or 13 or 14 or 15 or 16 or 17 or 18 or 19 or 20 or 21 or 22 or 23 or 24 or 25 or 26 or 27 or 28 or 29 or 30 or 31 or 32 or 33 or 34 or 35 or 36 or 37
39. Infectious Disease Transmission, Patient-to-Professional/
40. 38 and 39
41. exp Health Personnel/
42. (health care worker* or healthcare worker* or health care provider* or healthcare provider* or physiotherapist* or dentist* or nurse* or doctor* or physician* or health personnel or medical personnel or hospital personnel or hospital worker* or staff or healthcare professional* or health care professional* or care giver* or caregiver* or paramedic* or therapist*).ti,ab.
43. 41 or 42
44. Infectious Disease Transmission, Patient-to-Professional/
45. Occupational Exposure/
46. Air Microbiology/
47. Disease Transmission, Infectious/
48. infection control/ or infection control, dental/
49. exp Cross Infection/
50. Disease Outbreaks/
51. Aerosols/
52. ((aerosol* or cough* or droplet* or infection* or infectious or disease*) adj3 (generat* or induc* or stimulat* or produc*or creat* or respirable range* or dispers* or transmission or

transmitted or transmit or spread* or disseminat* or count* or precaution* or control* or inhibit* or prevent* or reduc*)).ti,ab.

53. cross-infection*.ti,ab.

54. 44 or 45 or 46 or 47 or 48 or 49 or 50 or 51 or 52 or 53

55. 38 and 43 and 54

56. (aerosol* adj2 generat* adj2 procedure*).ti,ab.

57. 40 or 55 or 56

58. Influenza, Human/

59. exp Influenza A virus/

60. SARS Virus/

61. Severe Acute Respiratory Syndrome/

62. exp Coronavirus/

63. Coronavirus Infections/

64. exp Tuberculosis/

65. exp Pneumonia/

66. Coronaviridae Infections/

67. (influenza* or H1N1 or tuberculosis or pneumonia or pneumococcus or severe acute respiratory syndrome or SARS or acute respiratory infection*).ti,ab.

68. (2019-nCoV* or 2019nCov* or coronavirus* or coronavirus 2 or coronavirus2* or corona or covid or covid-19 or covid19* or HCoV-19 or novel coronavirus* or ncov or SARS2 or SARS-COV-2* or SARS-COV2* or SARSCov or SARSCoV-2* or SARSCoV2* or Wuhan pneumonia or Wuhan virus).ti,ab,kf.

69. 58 or 59 or 60 or 61 or 62 or 63 or 64 or 65 or 66 or 67 or 68

70. 43 and 54 and 69

71. 57 or 70

72. limit 71 to (english or french)

73. limit 72 to yr="2011 -Current"

74. animals/ not humans/

75. 73 not 74

76. limit 75 to (editorial or letter)

77. 75 not 76

78. limit 77 to "review articles"

79. 77 not 78

80. limit 77 to "systematic review"

81. limit 77 to meta analysis

82. ((systematic or critical) adj1 (review* or overview* or synthesis)).ti,ab.

83. 77 and 82

84. 79 or 80 or 81 or 83

85. limit 84 to case reports

86. 84 not 85

87. limit 86 to comment

88. 86 not 87

**EMBASE Final March 31 2020**

1. positive end expiratory pressure/
2. exp High-Frequency Ventilation/
3. intermittent positive pressure ventilation/
4. endotracheal intubation/
5. endotracheal intubation/
6. Suction/
7. Tracheostomy/ or tracheobronchial toilet/
8. Bronchoscopy/
9. thorax drainage/
10. nebulization/ or exp nebulizer/
11. Sputum/ or sputum analysis/ or sputum examination/
12. Oxygen Therapy/
13. Autopsy/
14. exp lung function test/
15. exp lung function test/
16. resuscitation/
17. artificial ventilation/

18. exp Breathing Exercise/

19. (ventilation or ventilator or ventilating or ventilatory).ti,ab.

20. (respirator or respirators or respirat* support or respirat* care).ti,ab.

21. (intubation or intubated or extubation or extubated).ti,ab.

22. ((respiratory or airway or air way or open) adj3 suction*).ti,ab.

23. (nebulize* or nebulise* or aerosolize* or aerosolise*).ti,ab.

24. heat moisture exchange*.ti,ab.

25. (bronchoscopy or tracheostomy or thoracostomy).ti,ab.

26. (chest adj3 physiotherapy).ti,ab.

27. (sputum adj3 (induction or inducing)).ti,ab.

28. oxygen therap*.ti,ab.

29. (lung function test* or pulmonary function test*).ti,ab.

30. ((continuous or bilevel) adj2 (positive airway or positive pressure)).ti,ab.

31. (cardiopulmonary resuscitation or artificial resuscitation or artificial respiration).ti,ab.

32. (autopsy adj3 lung tissue*).ti,ab.

33. (bag adj1 mask*).ti,ab.

34. (ambu bag* or bag valve* or bvm).ti,ab.

35. (manual resuscitator* or self-inflating bag*).ti,ab.

36. ((nasopharyn* or nasal or nose or np or oropharyn* or OP) adj3 (culture* or smear* or specimen* or swab*)).ti,ab.

37. 1 or 2 or 3 or 4 or 5 or 6 or 7 or 8 or 9 or 10 or 11 or 12 or 13 or 14 or 15 or 16 or 17 or 18 or 19 or 20 or 21 or 22 or 23 or 24 or 25 or 26 or 27 or 28 or 29 or 30 or 31 or 32 or 33 or 34 or 35 or 36

38. exp Health Care Personnel/

39. (health care worker* or healthcare worker* or health care provider* or healthcare provider* or physiotherapist* or dentist* or nurse* or doctor* or physician* or health personnel or medical personnel or hospital personnel or hospital worker* or staff or healthcare professional* or health care professional* or care giver* or caregiver* or paramedic* or therapist*).ti,ab.

40. 38 or 39

41. *Occupational Exposure/

42. *airborne infection/ or *hospital infection/

43. *virus transmission/ or *bacterial transmission/
44. *infection control/
45. exp *Cross Infection/
46. *Disease transmission/
47. Aerosol/
48. ((aerosol* or cough* or droplet* or infection* or infectious or disease*) adj3 (generat* or induc* or stimulat* or produc*or creat* or respirable range* or dispers* or transmission or transmitted or transmit or spread* or disseminat* or count* or precaution* or control* or inhibit* or prevent* or reduc*)).ti,ab.
49. cross-infection*.ti,ab.
50. 41 or 42 or 43 or 44 or 45 or 46 or 47 or 48 or 49
51. 37 and 40 and 50
52. (aerosol* adj2 generat* adj2 procedure*).ti,ab.
53. 51 or 52
54. *Influenza/ or *influenza virus/
55. *Parainfluenza virus infection/
56. *Severe Acute Respiratory Syndrome/
57. *tuberculosis/ or *lung tuberculosis/ or *drug resistant tuberculosis/
58. *streptococcus pneumoniae/ or *pneumonia/ or *Respiratory syncytial pneumovirus/
59. exp *coronavirus infection/ or exp *coronavirinae/
60. (influenza* or H1N1 or tuberculosis or pneumonia or pneumococcus or severe acute respiratory syndrome or SARS or acute respiratory infection*).ti,ab.
61. (2019-nCoV* or 2019nCov* or coronavirus* or coronavirus 2 or coronavirus2* or corona or covid or covid-19 or covid19* or HCoV-19 or novel coronavirus* or ncov or SARS2 or SARS-COV-2* or SARS-COV2* or SARSCov or SARSCoV-2* or SARSCoV2* or Wuhan pneumonia or Wuhan virus).ti,ab,kw.
62. 54 or 55 or 56 or 57 or 58 or 59 or 60 or 61
63. 40 and 50 and 62
64. 53 or 63
65. limit 64 to (english or french)
66. limit 65 to yr="2011 -Current"

67. animals/ not human/

68. 66 not 67

69. limit 68 to conference abstract

70. 68 not 69

71. limit 70 to "review"

72. 70 not 71

73. limit 70 to (meta analysis or "systematic review")

74. ((systematic or crtical) adj1 (review* or overview* or synthesis)).tw.

75. 70 and 74

76. 72 or 73 or 75

77. case report/

78. 76 not 77

79. limit 78 to embase

**EBM Cochrane Database of Systematic Reviews March 31 2020**

1. (ventilation or ventilator or ventilating or ventilatory).ti,ab.

2. (respirator or respirators or respirat* support or respirat* care).ti,ab.

3. (intubation or intubated or extubation or extubated).ti,ab.

4. ((respiratory or airway or air way or open) adj3 suction*).ti,ab.

5. (nebulize* or nebulise* or aerosolize* or aerosolise*).ti,ab.

6. heat moisture exchange*.ti,ab.

7. (bronchoscopy or tracheostomy or thoracostomy).ti,ab.

8. (chest adj3 physiotherapy).ti,ab.

9. (sputum adj3 (induction or inducing)).ti,ab.

10. oxygen therap*.ti,ab.

11. (lung function test* or pulmonary function test*).ti,ab.

12. ((continuous or bilevel) adj2 (positive airway or positive pressure)).ti,ab.

13. (cardiopulmonary resuscitation or artificial resuscitation or artificial respiration).ti,ab.

14. (autopsy adj3 lung tissue*).ti,ab.

15. (bag adj1 mask*).ti,ab.

16. (ambu bag* or bag valve* or bvm).ti,ab.
17. (manual resuscitator* or self-inflating bag*).ti,ab.
18. ((nasopharyn* or nasal or nose or np or oropharyn* or OP) adj3 (culture* or smear* or specimen* or swab*)).ti,ab.
19. (health care worker* or healthcare worker* or health care provider* or healthcare provider* or physiotherapist* or dentist* or nurse* or doctor* or physician* or health personnel or medical personnel or hospital personnel or hospital worker* or staff or healthcare professional* or health care professional* or care giver* or caregiver* or paramedic* or therapist*).ti,ab.
20. ((aerosol* or cough* or droplet* or infection* or infectious or disease*) adj3 (generat* or induc* or stimulat* or produc*or creat* or respirable range* or dispers* or transmission or transmitted or transmit or spread* or disseminat* or count* or precaution* or control* or inhibit* or prevent* or reduc*)).ti,ab.
21. cross-infection*.ti,ab.
22. (aerosol* adj2 generat* adj2 procedure*).ti,ab.
23. (influenza* or H1N1 or tuberculosis or pneumonia or pneumococcus or severe acute respiratory syndrome or SARS or acute respiratory infection*).ti,ab.
24. (2019-nCoV* or 2019nCov* or coronavirus* or coronavirus 2 or coronavirus2* or corona or covid or covid-19 or covid19* or HCoV-19 or novel coronavirus* or ncov or SARS2 or SARS-COV-2* or SARS-COV2* or SARSCov or SARSCoV-2* or SARSCoV2* or Wuhan pneumonia or Wuhan virus).ti,ab.
25. 1 or 2 or 3 or 4 or 5 or 6 or 7 or 8 or 9 or 10 or 11 or 12 or 13 or 14 or 15 or 16 or 17 or 18
26. 20 or 21
27. 19 and 25 and 26
28. 22 or 27
29. 23 or 24
30. 19 and 26 and 29
31. 28 or 30

**EBM Cochrane CENTRAL Register March 31 2020**

1. exp Positive-Pressure Respiration/

2. exp High-Frequency Ventilation/
3. exp Ventilators, Mechanical/
4. Ventilation/
5. exp Intubation, Intratracheal/
6. Suction/
7. Tracheostomy/
8. Bronchoscopy/
9. Thoracostomy/
10. exp "Nebulizers and Vaporizers"/
11. Sputum/
12. Oxygen Inhalation Therapy/
13. Autopsy/
14. exp Respiratory Function Tests/
15. exp Spirometry/
16. exp Cardiopulmonary Resuscitation/
17. exp Respiration, Artificial/
18. exp Breathing Exercises/
19. Physical Therapy Modalities/ and Thorax/
20. (ventilation or ventilator or ventilating or ventilatory).ti,ab.
21. (respirator or respirators or respirat* support or respirat* care).ti,ab.
22. (intubation or intubated or extubation or extubated).ti,ab.
23. ((respiratory or airway or air way or open) adj3 suction*).ti,ab.
24. (nebulize* or nebulise* or aerosolize* or aerosolise*).ti,ab.
25. heat moisture exchange*.ti,ab.
26. (bronchoscopy or tracheostomy or thoracostomy).ti,ab.
27. (chest adj3 physiotherapy).ti,ab.
28. (sputum adj3 (induction or inducing)).ti,ab.
29. oxygen therap*.ti,ab.
30. (lung function test* or pulmonary function test*).ti,ab.
31. ((continuous or bilevel) adj2 (positive airway or positive pressure)).ti,ab.
32. (cardiopulmonary resuscitation or artificial resuscitation or artificial respiration).ti,ab.

33. (autopsy adj3 lung tissue*).ti,ab.
34. (bag adj1 mask*).ti,ab.
35. (ambu bag* or bag valve* or bvm).ti,ab.
36. (manual resuscitator* or self-inflating bag*).ti,ab.
37. ((nasopharyn* or nasal or nose or np or oropharyn* or OP) adj3 (culture* or smear* or specimen* or swab*)).ti,ab.
38. 1 or 2 or 3 or 4 or 5 or 6 or 7 or 8 or 9 or 10 or 11 or 12 or 13 or 14 or 15 or 16 or 17 or 18 or 19 or 20 or 21 or 22 or 23 or 24 or 25 or 26 or 27 or 28 or 29 or 30 or 31 or 32 or 33 or 34 or 35 or 36 or 37
39. Infectious Disease Transmission, Patient-to-Professional/
40. 38 and 39
41. exp Health Personnel/
42. (health care worker* or healthcare worker* or health care provider* or healthcare provider* or physiotherapist* or dentist* or nurse* or doctor* or physician* or health personnel or medical personnel or hospital personnel or hospital worker* or staff or healthcare professional* or health care professional* or care giver* or caregiver* or paramedic* or therapist*).ti,ab.
43. 41 or 42
44. Infectious Disease Transmission, Patient-to-Professional/
45. Occupational Exposure/
46. Air Microbiology/
47. Disease Transmission, Infectious/
48. infection control/ or infection control, dental/
49. exp Cross Infection/
50. Disease Outbreaks/
51. Aerosols/
52. ((aerosol* or cough* or droplet* or infection* or infectious or disease*) adj3 (generat* or induc* or stimulat* or produc*or creat* or respirable range* or dispers* or transmission or transmitted or transmit or spread* or disseminat* or count* or precaution* or control* or inhibit* or prevent* or reduc*)).ti,ab.
53. cross-infection*.ti,ab.
54. 44 or 45 or 46 or 47 or 48 or 49 or 50 or 51 or 52 or 53

55. 38 and 43 and 54

56. (aerosol* adj2 generat* adj2 procedure*).ti,ab.

57. 40 or 55 or 56

58. Influenza, Human/

59. exp Influenza A virus/

60. SARS Virus/

61. Severe Acute Respiratory Syndrome/

62. exp Coronavirus/

63. Coronavirus Infections/

64. exp Tuberculosis/

65. exp Pneumonia/

66. Coronaviridae Infections/

67. (influenza* or H1N1 or tuberculosis or pneumonia or pneumococcus or severe acute respiratory syndrome or SARS or acute respiratory infection*).ti,ab.

68. (2019-nCoV* or 2019nCov* or coronavirus* or coronavirus 2 or coronavirus2* or corona or covid or covid-19 or covid19* or HCoV-19 or novel coronavirus* or ncov or SARS2 or SARS-COV-2* or SARS-COV2* or SARSCov or SARSCoV-2* or SARSCoV2* or Wuhan pneumonia or Wuhan virus).ti,ab.

69. 58 or 59 or 60 or 61 or 62 or 63 or 64 or 65 or 66 or 67 or 68

70. 43 and 54 and 69

71. 57 or 70

72. limit 71 to (english or french)

73. limit 72 to yr="2011 -Current"

74. animals/ not humans/

75. 73 not 74

**CINAHL Search March 31 2020**

1. ((MH "Positive Pressure Ventilation") OR (MH "Intermittent Positive Pressure Ventilation") OR (MH "Continuous Positive Airway Pressure") OR (MH "Ventilation, High Frequency") OR (MH "Ventilators, Mechanical") OR (MH "Ventilation, Mechanical, Differentiated") OR

(MH "Ventilation") OR (MH "Suction") OR (MH "Tracheostomy") OR (MH "Bronchoscopy") OR (MH "Thoracostomy") OR (MH "Nebulizers and Vaporizers") OR (MH "Sputum") OR (MH "Oxygen Therapy") OR (MH "Autopsy") OR (MH "Respiratory Function Tests+") OR (MH "Spirometry") OR (MH "Resuscitation, Cardiopulmonary") OR (MH "Respiration, Artificial") OR (MH "Breath Tests") OR (MH "Breathing Exercises") ) OR TI ( Ventilat* or respirat* or intubat* or extubat* or nebulize* or nebulise* or aerosolize* or aerosolise* or heat moisture exchange* or bronchoscopy or tracheostomy or thoracostomy or oxygen therap* or lung function test* or pulmonary function test* or cardiopulmonary resuscitation or artificial resuscitation or artificial respiration or ambu bag* or bag valve* or bvm or manual resuscitator* or self-inflating bag* ) OR AB ( Ventilat* or respirat* or intubat* or extubat* or nebulize* or nebulise* or aerosolize* or aerosolise* or heat moisture exchange* or bronchoscopy or tracheostomy or thoracostomy or oxygen therap* or lung function test* or pulmonary function test* or cardiopulmonary resuscitation or artificial resuscitation or artificial respiration or ambu bag* or bag valve* or bvm or manual resuscitator* or self-inflating bag* )

2. TI ( (nasopharyn* or nasal or nose or np or oropharyn* or OP) N3 (culture* or smear* or specimen* or swab*) ) OR AB ( (nasopharyn* or nasal or nose or np or oropharyn* or OP) N3 (culture* or smear* or specimen* or swab*) )
3. TI ( (respiratory or airway or air way or open) N3 suction* ) OR AB ( (respiratory or airway or air way or o[illegible]
5. TI chest N3 physiotherapy OR AB chest N3 physiotherapy
6. TI ( sputum N3 (induction or inducing) ) OR AB ( sputum N3 (induction or inducing) )
7. TI ( (continuous or bilevel) N2 (positive airway or positive pressure) ) OR AB ( (continuous or bilevel) N2 (positive airway or positive pressure) )
8. TI autopsy N3 lung tissue* OR AB autopsy N3 lung tissue*
9. TI bag n1 mask* OR AB bag n1 mask*
10. 1 or 2 or 3 or 4 or 5 or 6 or 7 or 8 or 9
11. (MH "Disease Transmission, Patient-to-Professional")
12. 10 and 11
13. MH "Health Personnel+") OR TI ( (health care worker* or healthcare worker* or health care provider* or healthcare provider* or physiotherapist* or dentist* or nurse* or doctor* or physician* or health personnel or medical personnel or hospital personnel or hospital

worker* or staff or healthcare professional* or health care professional* or care giver* or caregiver* or paramedic* or therapist*) ) OR AB ( (health care worker* or healthcare worker* or health care provider* or healthcare provider* or physiotherapist* or dentist* or nurse* or doctor* or physician* or health personnel or medical personnel or hospital personnel or hospital worker* or staff or healthcare professional* or health care professional* or care giver* or caregiver* or paramedic* or therapist*) )

14. (MH "Occupational Exposure") OR (MH "Air Microbiology") OR (MH "Disease Transmission, Patient-to-Professional") OR (MH "Infection Control") OR (MH "Disease Transmission") OR (MH "Cross Infection") OR (MH "Disease Outbreaks") ) OR TI ( ((aerosol* or cough* or droplet* or infection* or infectious or disease*) N3 (generat* or induc* or stimulat* or produc*or creat* or respirable range* or dispers* or transmission or transmitted or transmit or spread* or disseminat* or count* or precaution* or control* or inhibit* or prevent* or reduc*)) ) OR AB ( ((aerosol* or cough* or droplet* or infection* or infectious or disease*) N3 (generat* or induc* or stimulat* or produc*or creat* or respirable range* or dispers* or transmission or transmitted or transmit or spread* or disseminat* or count* or precaution* or control* or inhibit* or prevent* or reduc*)) ) OR TI cross-infection* OR AB cross-infection*
15. 10 and 13 and 14
16. 12 or 15
17. TI aerosol* N2 generat* N2 procedure* OR AB aerosol* N2 generat* N2 procedure*
18. 16 or 17
19. (MH "Influenza, Human") OR (MH "Influenza A H5N1") OR (MH "Influenza A Virus") OR (MH "Influenza, Pandemic (H1N1) 2009") OR (MH "Influenza, Seasonal") OR (MH "SARS Virus") OR (MH "Influenza A Virus, H1N1 Subtype") OR (MH "Influenza A Virus, H5N1 Subtype") OR (MH "Severe Acute Respiratory Syndrome") OR (MH "Respiratory Distress Syndrome, Acute") OR (MH "Respiratory Distress Syndrome") OR (MH "Coronavirus") OR (MH "Coronavirus Infections") OR (MH "Tuberculosis") OR (MH "Pneumonia+") OR (MH "Coronaviridae Infections") ) OR TI ( (influenza* or H1N1 or tuberculosis or pneumonia or pneumococcus or severe acute respiratory syndrome or SARS or acute respiratory infection*) ) OR AB ( (influenza* or H1N1 or tuberculosis or pneumonia or pneumococcus or severe acute respiratory syndrome or SARS or acute respiratory infection*) ) OR TI ( (2019-nCoV*

or 2019nCov* or coronavirus* or coronavirus 2 or coronavirus2* or corona or covid or covid-19 or covid19* or HCoV-19 or novel coronavirus* or ncov or SARS2 or SARS-COV-2* or SARS-COV2* or SARSCov or SARSCoV-2* or SARSCoV2* or Wuhan pneumonia or Wuhan virus) ) OR AB ( (2019-nCoV* or 2019nCov* or coronavirus* or coronavirus 2 or coronavirus2* or corona or covid or covid-19 or covid19* or HCoV-19 or novel coronavirus* or ncov or SARS2 or SARS-COV-2* or SARS-COV2* or SARSCov or SARSCoV-2* or SARSCoV2* or Wuhan pneumonia or Wuhan virus) )

20. 13 and 18 and 19
21. 18 or 20
22. **Limiters** - Published Date: 20110101-20201231; Scholarly (Peer Reviewed) Journals
23. **Narrow by Language:** - English
