## Supplementary material for "Transmission of Acute Respiratory Infections during Aerosol Generating Medical Procedures (AGMPs): An updated review": A3 Appendix

**Additional File 3. Table of Excluded Articles**

| Author (Year) | Reason for Exclusion |
|---|---|
| Anonymous (2011)[1] | Other |
| Baig et al. (2015)[2] | Study design not of interest |
| Bhadelia et al. (2013)[3] | Study design not of interest |
| Bien et al. (2016)[4] | Study design not of interest |
| Bischoff et al. (2013)[5] | Intervention not of interest |
| Chan et al. (2018)[6] | Comparator not of interest |
| Chin et al. (2014)[7] | Outcome not of interest |
| Chughtai et al. (2016)[8] | Outcome not of interest |
| Chung et al. (2015)[9] | Outcome not of interest |
| Cummings et al. (2014)[10] | Study design not of interest |
| De Perio et al. (2014)[11] | Intervention not of interest |
| De Vries et al. (2017)[12] | Intervention not of interest |
| Du et al. (2012)[13] | Intervention not of interest |
| Hui et al. (2013)[14] | Comparator not of interest |
| Hui et al. (2014)[15] | Intervention not of interest |
| Ito et al. (2016)[16] | Intervention not of interest |
| Jones et al. (2018)[17] | Intervention not of interest |
| Kim et al. (2016)[18] | Comparator not of interest |
| Kim et al. (2015)[19] | Outcome not of interest |
| Kim et al. (2015)[20] | Outcome not of interest |
| Kim et al. (2013)[21] | Intervention not of interest |
| King et al. (2015)[22] | Intervention not of interest |
| Kulkarni et al. (2016)[23] | Other |
| Lee et al. (2015)[24] | Study design not of interest |
| MacIntyre et al. (2017)[25] | Intervention not of interest |
| MacIntyre et al. (2014)[26] | Study design not of interest |
| Memish et al. (2018)[27] | Intervention not of interest |
| Mitchell et al. (2012)[28] | Outcome not of interest |
| Moon et al. (2019)[29] | Comparator not of interest |
| Morilla et al. (2018)[30] | Comparator not of interest |
| Na et al. (2016)[31] | Outcome not of interest |
| Nam et al. (2017)[32] | Comparator not of interest |
| O'Neil et al. (2017)[33] | Other |
| Offeddu et al. (2017)[34] | Intervention not of interest |
| Pedrosa et al. (2011)[35] | Intervention not of interest |
| Ricard et al. (2011)[36] | Comparator not of interest |
| Rule et al. (2018)[37] | Outcome not of interest |
| Sayed et al. (2011)[38] | Intervention not of interest |
| Schmidt et al. (2018)[39] | Intervention not of interest |

| | |
|---|---|
| Schubert et al. (2019)[40] | Intervention not of interest |
| Schwarz et al. (2019)[41] | Intervention not of interest |
| Seto (2015)[42] | Study design not of interest |
| Seto et al. (2013)[43] | Study design not of interest |
| Seto et al. (2011)[44] | Intervention not of interest |
| Shin et al. (2017)[45] | Outcome not of interest |
| Shiu et al. (2020)[46] | Intervention not of interest |
| Suen et al. (2017)[47] | Outcome not of interest |
| Thompson et al. (2013)[48] | Outcome not of interest |
| Tran et al. (2013)[49] | Other |
| Tran et al. (2012)[50] | Other |
| Tsai et al. (2015)[51] | Outcome not of interest |
| Tudor et al. (2014)[52] | Intervention not of interest |
| Uden et al. (2017)[53] | Intervention not of interest |
| Weber et al. (2019)[54] | Outcome not of interest |
| Zietsman et al. (2019)[55] | Comparator not of interest |
| Zumla et al. (2014)[56] | Comparator not of interest |
| Zuo et al. (2020)[57] | Comparator not of interest |


1. Anonymous. Flu outbreak points to risk from ill co-workers. Relias Media: Continuing Education. 2011.
2. Baig S, Rashid T, Saleem M. Protection from blood aerosol contamination when managing epistaxis: A study of the effeciveness of a patient mouth mask. ENT Journal. 2015.
3. Bhadelia N, Sonti R, McCarthy JW, Vorenkamp J, Jia H, Saiman L, et al. Impact of the 2009 Influenza A (H1N1) Pandemic on Healthcare Workers at a Tertiary Care Center in New York City. Infect Control Hosp Epidemiol. 2013;34(8):825-31.
4. Bien EA, Gillespie G, Betcher C, Thrasher T, Mingerink D. Respiratory protection toolkit: providing guidance without changing requirements - can we make an impact? Workplace Health Safety. 2016.
5. Bischoff WE, Swett K, Leng I, Peters TR. Exposure to Influenza Virus Aerosols During Routine Patient Care. The Journal of Infectious Diseases. 2013;207(7):1037-46.
6. Chan MTV, Chow BK, Lo T, Ko FW, Ng SS, Gin T, et al. Exhaled air dispersion during bag-mask ventilation and sputum suctioning - Implications for infection control. Sci Rep. 2018;8(1):198.
7. Chin TL, MacGowan AP, Jacobson SK, Donati M. Viral infections in pregnancy: advice for healthcare workers. Journal of Hospital Infection. 2014;87(1):11-24.
8. Chughtai AA, Seale H, Dung TC, Hayen A, Rahman B, Raina MacIntyre C. Compliance with the Use of Medical and Cloth Masks Among Healthcare Workers in Vietnam. ANNHYG. 2016;60(5):619-30.
9. Chung FF, Lin HL, Liu HE, Lien ASY, Hsiao HF, Chou LT, et al. Aerosol Distribution During Open Suctioning and Long-Term Surveillance of Air Quality in a Respiratory Care Center Within a Medical Center. Respiratory Care. 2015;60(1):30-7.

26. MacIntyre CR, Richards GA, Davidson PM. Respiratory protection for healthcare workers treating Ebola virus disease (EVD): Are facemasks suficient to meet occupational health and safety obligations? International Journal of Nursing Studies. 2014;51.
27. Memish ZA, Perlman S, Van Kerkhove MD, Zumla A. Middle east respiratory syndrome. Lancet. 2018;395.
28. Mitchell R, Ogunremi T, Astrakianakis G, Bryce E, Gervais R, Gravel D, et al. Impact of the 2009 influenza A (H1N1) pandemic on Canadian health care workers: A survey on vaccination, illness, absenteeism, and personal protective equipment. American Journal of Infection Control. 2012;40(7):611-6.
29. Moon J, Lee H, Jeon JH, Kwon Y, Kim H, Wang EB, et al. Aerosol transmission of severe fever with thrombocytopenia syndrome virus during resuscitation. Infect Control Hosp Epidemiol. 2019;40(2):238-41.
30. Morilla R, Martinez MT, de la Horra C, Frizaz V, Martin-Juan JM, Romero B, et al. Airborne acquisition of Pneumocystis in bronchoscopy units: a hidden danger to healthcare workers. International society for human and animal mycology. 2019.
31. Na HJ, Eom JS, Lee G, Mok JH, Kim MH, Lee KS, et al. Exposure to myobacterium tuberculosis during flexible bronchoscopy in patients with unexpected pulmonary tuberculosis. PLoS ONE. 2016.
32. Nam HS, Yeon MY, Park JW, Hong JY, Song JW. Healthcare worker infected with midle east respiratory syndrome during cardiopulmonary resuscitation in Korea. Epidemiology and Health. 2017;39.
33. O'Neil CA, Li J, Leavey A, Wang Y, Hink M, Wallace M, et al. Characterization of Aerosols Generated During Patient Care Activities. Clinical Infectious Diseases. 2017;65(8):1342-8.
34. Offeddu V, Yung CF, Low MSF, Tam CC. Effectiveness of Masks and Respirators Against Respiratory Infections in Healthcare Workers: A Systematic Review and Meta-Analysis. Clinical Infectious Diseases. 2017;65(11):1934-42.
35. Pedrosa PBS, Cardoso TAO. Viral infections in workers in hospital and research laboratory settings: a comparative review of infection modes and respective biosafety aspects. International Journal of Infectious Diseases. 2011;15(6):e366-e76.
36. Ricard J-D, Eveillard M, Martin Y, Barnaud G, Branger C, Dreyfuss D. Influence of tracheal suctioning systems on health care workers' gloves and equipment contamination: A comparison of closed and open systems. American Journal of Infection Control. 2011;39(7):605-7.
37. Rule AM, Apau O, Ahrenholz SH, Brueck SE, Lindsley WG, de Perio MA, et al. Healthcare personnel exposure in an emergency department during influenza season. PLoS ONE. 2018;13(8):e0203223.
38. Sayed ME, Kue R, McNeil C, Dyer KS. A Descriptive Analysis of Occupational Health Exposures in an Urban Emergency Medical Services System: 2007–2009. Prehospital Emergency Care. 2011;15(4):506-10.
39. Schmidt B-M, Engel ME, Abdullahi L, Ehrlich R. Effectiveness of control measures to prevent occupational tuberculosis infection in health care workers: a systematic review. BMC Public Health. 2018;18(1):661.
40. Schubert M, Kampf D, Jatzwauk L, Kynast F, Stein A, Strasser R, et al. Prevalence and predictors of MRSA carriage among employees in a non-outbreak setting: a cross-sectional study in an acute care hospital. Journal of Occupational Medicine and Toxicology. 2019.

41. Schwartz H, Boni J, Kouyos RD, Turk T, Battegay E, Kohler M, et al. The TransFLUas influenza transmission study in acute healthcare - recruitment rates and protocol adherence in healthcare workers and inpatients. BMC Infectious Diseases. 2019.
42. Seto WH. Airborne transmission and precautions: facts and myths. Journal of Hospital Infection. 2015;89.
43. Seto WH, Conly JM, Pessoa-Silva CL, Malik M, Eremin S. Infection prevention and control measures for acute respiratory infections in healthcare settings: an update. Eastern Mediterranean Health Journal. 2013;19.
44. Seto WH, Cowling BJ, Lam H, Ching PTY, To ML, Pittet D. Clinical and nonclinical health care workers faced a similar risk of acquiring 2009 pandemic H1N1 infection. CID. 2011;53.
45. Shin H, Oh J, Lim TH, Kang H, Song Y, Lee S. Comparing the protective performances of 3 types of N95 filtering facepiece respirators during chest compressions. Medicine. 2017;96(42).
46. Shiu EYC, Huang W, Ye D, Xie Y, Mo J, Li Y, et al. Frequent recovery of influenza A but not influenza B virus RNA in aerosols in pediatric patient rooms. Indoor Air. 2020.
47. Suen LKP, Yang L, Fung KHK, Boost MV, Wu CST, Au-Yeung C, et al. Reliability of N95 repirators for respiratory protection before, during, and after nursing procedures. American Journal of Infection Control. 2017;45.
48. Thompson KA, Pappachan JV, Bennett AM, Mittal H, Macken S, Dove BK, et al. Inflenza aerosols in UK Hospitals during the H1N1 (2009) pandemic - the risk of aerosol generation during medical procedures. PLoS ONE. 2013;8.
49. Tran K, Cimon K, Severn M, Pessoa-sSilva CL, Conly J. Aerosol-generating procedures and risk of transmission of acute respiratory infections: a systematic review. CADTH Technology Overviews. 2013 3(1).
50. Tran K, Cimon K, Severn M, Pessoa-Silva CL, Conly J. Aerosol generating procedures and risk of transmission of acute respiratory infections to healthcare workers: a systematic review. PLoS ONE. 2012.
51. Tsai RJ, Boiano JM, Steege AL, Sweeney MH. Precautionary practices of respiratory therapists and other health-care practitioners who administer aerosolized medications. Respiratory Care. 2015;60.
52. Tudor C, Van der Walt M, Margot B, Dorman SE, Pan WK, Yenokyan G, et al. Tuberculosis among health care workers in KwaZulu-Natal, South Africa: a retrospective cohort analysis. BMC Public Health. 2014;14(891).
53. Uden L, Barber E, Ford N, Cooke GS. Risk of tuberculosis infection and disease for health care workers: an updated meta-analysis. Open Forum Infection Diseases. 2017.
54. Weber RT, Phan LT, Fritzen-Pedicini C, Johnes RM. Environmental and personal protective equipment contamination during simulated healthcare activities. Annals of work exposures and health. 2019;63.
55. Zietsman M, Phan LT, Jones RM. Potential for occupational exposures to pathogens during bronchoscopy procedures. Journal of Occupational and Environmental Hygiene. 2019;16.
56. Zumla A, Hui DSC. Infection control and MERS-CoV in health-care workers. Lancet. 2014;383.
57. Zuo M, Huang Y, Ma W, Xue Z, Zhang J, Gong Y, et al. Expert recommendations for tracheal intubation in critically ill patients with noval coronavirus disease 2019. Chinese medical sciences journal. 2020.
